# Real-world performance and safety of Cerviron® medical device in the treatment of various types of vaginitis

**DOI:** 10.1101/2023.05.11.22282141

**Authors:** Izabella Petre, Daniela Teodora Sirbu, Ramona Petrita, Florentina Liliana Calancea, Andreea-Denisa Toma, Ema Peta, Florentina Dimcevici-Poesina

## Abstract

A collection of clinical data was conducted to assess the performance and tolerability of Cerviron^®^ ovules in the treatment and management of various types of vaginitis in clinical practice. A total of 111 women aged between 20 and 70 years were included, 71 of whom were treated with Cerviron^®^ ovules as monotherapy and 40 who used Cerviron^®^ ovules as supportive treatment in conjunction with antibiotic therapy. The aim of our study was to assess the relief in vaginal symptoms and changes in the normal vaginal pH level after 3 months of treatment with Cerviron^®^ medical device in real-life clinical practice settings. The results showed that Cerviron^®^ ovules are well tolerated and effective as monotherapy and also as an adjuvant to antibiotic therapy. The study and its details are registered in www.clinicaltrials.gov under ID NCT05652959.

## INTRODUCTION

Vaginitis is a broad term that includes a range of gynecological disorders characterized by infection of vaginal mucosa, inflammation of vulva, and alteration of the normal vaginal microflora. Endogenous or/and exogenous microorganisms may cause various vaginal symptoms such as abnormal leucorrhea, pain, dysuria, dyspareunia, local edema and redness. Moreover, the physiological changes in vaginal climate influenced by vaginitis are correlated with the susceptibility of acquiring HIV infection and other sexually transmitted infections. (1) The most common infectious forms of vaginitis include bacterial vaginosis, vulvovaginal candidiasis, and trichomoniasis. The endogenous infections (named non-specific vaginitis) are caused by the normal vaginal flora, such as anaerobic gram-negative bacilli, anaerobic gram-negative and gram-positive cocci. (2) Currently, the anti-infectious therapy is usually prescribed in infectious vaginitis or mixed vaginitis. However, antibiotic treatment also affects vaginal Lactobacilli, that protect the vaginal microenvironment and can cause vulvovaginal mycotic infections, frequent relapse of vaginitis or other side effects such as headache, nausea, diarrhea, dysmenorrhea or vulvovaginal pruritus. (3)

Relapsing vaginal symptoms negatively affect patients ‘quality of life in terms of discomfort and pain, professional activity, sexual functioning, and self-image. (4) Co-infections are common in vaginitis, making accurate diagnosis and treatment very challenging, due to the fact that the polymicrobial interactions and mixed biofilms cause therapeutic failure, recurrence and reinfection. More than 20% of infectious vaginitis cases may be mixed. (5,6) In the past years, this challenging gynecological disorder has all too often been ignored by the medical community or regarded merely as a minor annoyance to women an therefore no standard of care is yet defined for any type of vaginitis. (7,8)

Bacterial vaginosis was reported in literature as the most prevalent vaginal infection, with an approximate 50 % of the total cases of vaginal infections. (9) Most of the symptomatic patients self-medicate before seeking an evaluation by a medical professional, because complementary, alternative therapies and over-the-counter medications are available in a varied range. (10,11) Prevention of infection should include avoiding long-term use of antibiotics and a clear contraindication to frequent vaginal douching.(12) Recurrence of vaginosis is really frequent, and can include four or more episodes in one year. (13–15)

Non-infectious vaginitis (such as atrophic, irritant, and inflammatory vaginitis) are less common and account for less than 10 % of the total cases of vaginal infections. (8,16) Irritant/allergic vaginitis is caused by allergies to vaginal sprays, douches, spermicides, soaps, detergents or fabric softeners. These products can cause specific vaginal symptoms such as burning, itching, and discharge, even if there is no infection. Vaginal irritation (or vaginal atrophy) caused by the natural lessening in estrogen levels can appear during breast-feeding and after menopause. (17)

## MATERIALS AND METHODS

Cerviron^®^ is a medical device manufactured by PFC Pharma Manufacturing SL and formulated following the provisions of the European Regulation 2017/745 on Medical Devices. Cerviron^®^ is an invasive medical device of short-term use classified under annex VIII of the European Regulation 2017/745 as class IIb according to Rule 21 (devices composed of substances or of combinations of substances to be introduced into the human body via a body orifice or applied to the skin and absorbed by or locally dispersed in the human body). Cerviron^®^ has a complex composition consisting of three topical pharmaceutical products – hexylresorcinol, collagen and bismuth subgallate – and four phytotherapeutic extracts – Calendula officinalis, Hydrastis canadensis, Thymus vulgaris extract and Curcuma longa. The Instructions for Use specify its field of use as adjuvant in the treatment of acute and chronic vulvovaginitis of mechanical etiology, caused by changes of vaginal pH and changes of the vaginal flora and of cervical lesions of mechanical origin.

Cerviron^®^ ovules is a CE-mark medical device available on European and Middle-East markets in the following countries: Albania, Latvia, Lithuania, Estonia, Kosovo, Montenegro, Romania, Kuwait, and United Arab Emirates. This medical device has a local therapeutic action, with indication in the treatment of atrophic, aerobic and traumatic vaginitis, as well as in the treatment of cervical lesions.

The present investigation was designed as real-world evidence study with the primary purpose of assessing the performance and tolerance of Cerviron^®^ ovules in the treatment and management of various types of vulvovaginitis. The study collected clinical data from 28 different specialized gynecology clinical facilities. The study was conducted between May 20, 2021 and August 31, 2021.

The primary objective of this study was to assess the tolerability of Cerviron^®^ ovules in the management of various types of vulvovaginitis, but also to confirm its performance both on symptoms relief and as a user-friendly device. The secondary objective of this study was to assess the performance of the medical device by clinical exam and patients’ degree of satisfaction. Medical charts of patients under treatment with Cerviron^®^ for at least three months were analyzed.

### Study population

Data of 111 women aged between 20 and 70 years were analyzed, 71 of whom were treated with Cerviron® ovules as monotherapy and 40 who used Cerviron® ovules as supportive treatment in conjunction with the antibiotic therapy.

The following symptoms were recorded in the medical charts: leukorrhea, vaginal itching, pain and a feeling of tension, vaginal burning sensation, erythema, abnormal odor of vaginal secretions, dysuria and dyspareunia. The prevalence rates of vulvovaginitis and bacterial vaginosis were 89.191% (99/111) and 10.81% (12/111), respectively.

Each specialist decided, in a case-by-case situation, to prescribe Cerviron^®^ as a sole treatment or in combination with anti-infectious therapy. Thus, 71 participants were treated with Cerviron® monotherapy and 40 participants were treated with a combination of antibiotics and Cerviron® ovules. The inclusion criteria included participants with a diagnosis of vaginitis and specific symptomatology.

This investigation was conducted in accordance with the Medical Devices Coordination Group MDCG 2020-7 Post-market clinical follow-up requirements, Medical Devices Regulation (EU) 2017/745 and Study of medical devices for human subjects — Good clinical practice, ISO 14155:2020.

### Statistical methods

Quantitative variables (i.e., demographics) were expressed as mean ± standard deviation (SD) if normally distributed, otherwise median, minimum, maximum and interquartile range were given. Qualitative variables were scored using frequencies and percentages. To assess changes over time before and after treatment, we performed Paired t-tests (where applicable) or Wilcoxon signed rank sums for quantitative variables and used McNemar’s test to assess changes in binary variables.

The quality and completeness of the collected data were preliminarily assessed in comparison with data analysis. If medical charts recorded no information for one or more variables, the missing data was not replaced.

Data analysis was performed after the first, second and third treatment levels. Safety evaluations included intention-to-treat (ITT) cohort consisting of all participants who received Cerviron^®^ for at least three months. Efficacy analyzes were performed in the Per Protocol (PP) population, which included only participants who attended the third follow-up visit (3 months).

## RESULTS

Primary objective included number of possible adverse reactions observed during the treatment. Secondary performance objectives included the improvement in the vaginal discharge aspect (normal/abnormal), presence/absence of burn and/or vaginal pain and bad odour, presence/absence of vaginal irritation and changed in vaginal pH values.

In order to calculate the statistical significance related to the performance of the medical device, it was used a one-tailed z-test (0.05 significance level) with the first screening visit as baseline (null hypothesis that is no statistically significant difference between baseline visit and other treatment visits). The following p-values resulted (calculated for each of the following visits, to evaluate the significance at 30, 60 and 90 days of treatment), as seen in Table 1.

**Table 1.** p-values to evaluate the significance at 30, 60 and 90 days of treatment) *Note: For the p-value calculation, an improvement or good performance of the indicator was considered if the doctor rated the result with “Good” or “Very Good.

| No of days | Abnormal vaginal discharge | Burns and/ or vaginal pain | Abnormal smell | Changes in the normal pH level |
| --- | --- | --- | --- | --- |
| 30 days | $p < 0.0001$ | $p < 0.0001$ | $p < 0.0001$ | $p < 0.0001$ |
| 60 days | $p < 0.0001$ | $p < 0.0001$ | $p < 0.0001$ | $p < 0.0001$ |
| 90 days | $p < 0.0001$ | $p < 0.0001$ | $p < 0.0001$ | $p < 0.0001$ |

The overall indicators of the study are presented below.

### 1. Abnormal vaginal discharge

Assessment of abnormal vaginal discharge was performed for 100 study participants, and was rated as follows: absent in 20%, mild in 50%, and moderate in 30% of the evaluated participants. Out of 91 patients examined, 50.55% had normal discharge and in 42.86% a mild abnormal discharge was recorded. As seen in figure 1, the final visit showed the following results: 89.53% had no abnormal discharge, 8.14% had mild discharge, and only 1.16% had moderate or similar abnormal discharge before treatment.

**Figure 1.**
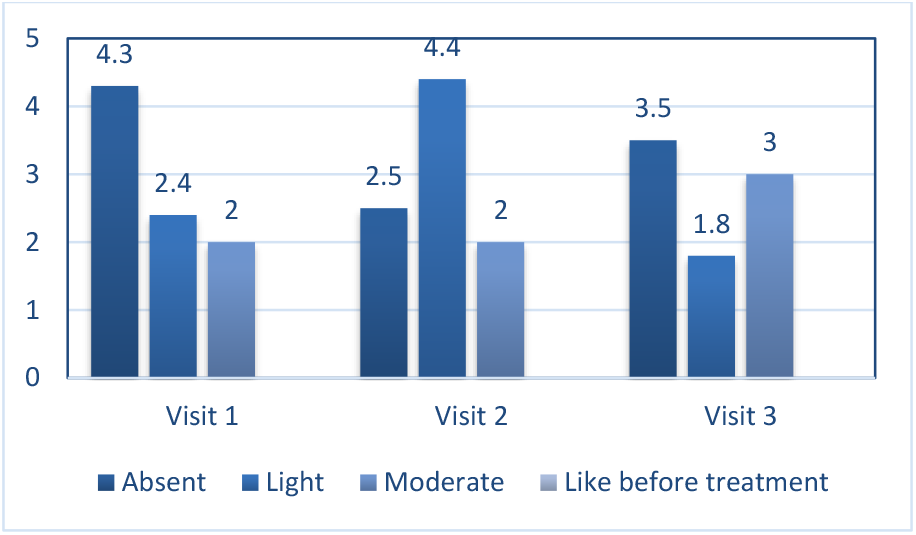
Abnormal vaginal discharge evaluation at every visit

When using Cerviron® ovules as monotherapy, in termen of vaginal symptoms relief, the results show that 80.85% were asymptomatic, 10.64% had mild changes, 8.51% had moderate changes, and none of the patients had the same symptoms as before treatment. The vaginal discharge change was evaluated in the participants treated with monotherapy vs Cerviron® with additional anti- infectious therapies, as shown in figure 2.

**Figure 2.**
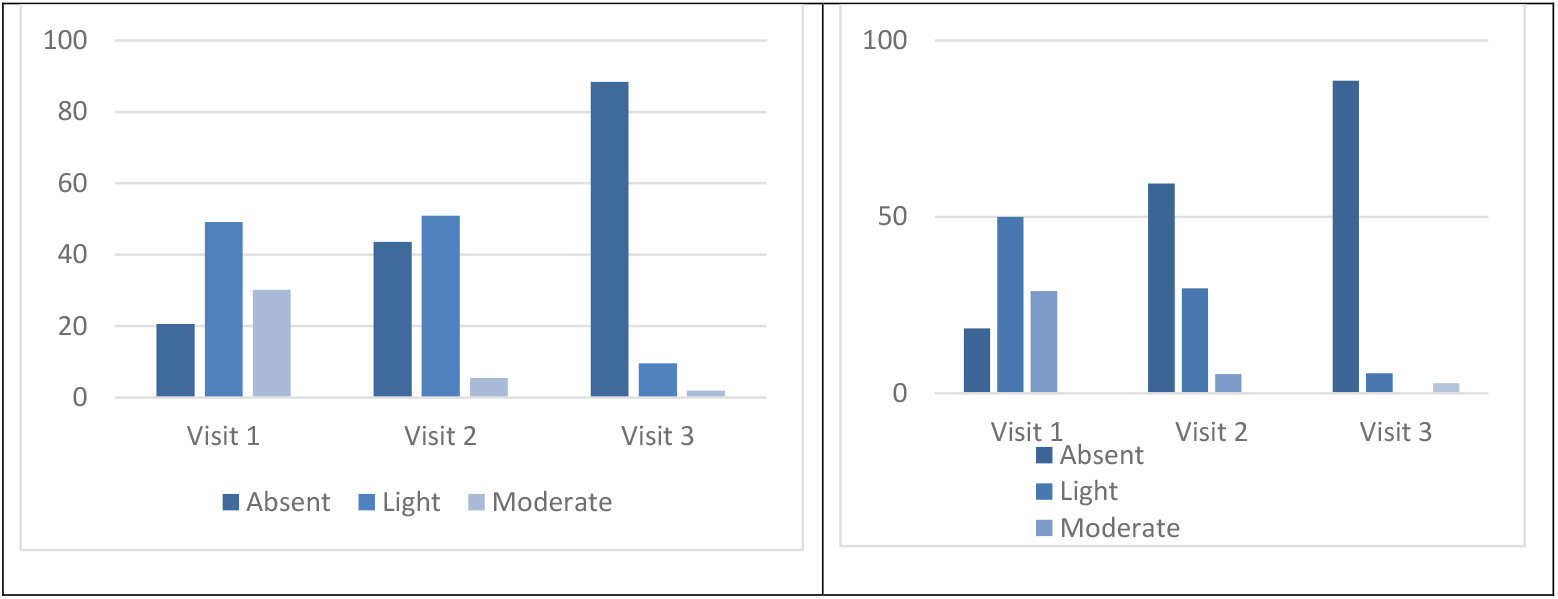
Performance of Cerviron® ovules as monotherapy (left) vs Cerviron® ovules as adjuvant treatment (right): in treating abnormal vaginal discharge

In monotherapy, out of 55 patients evaluated at the second visit, 43.64% had normal vaginal discharge, and 50.91% assessed the symptom as light abnormal discharge. At Visit 3, 88.46% participants had a normal vaginal discharge, 9.62% had mild vaginal discharge and only 1.92% had moderate abnormal vaginal discharge. As adjuvant treatment, Cerviron^®^ ovules performed well, at Visit 88.57% participants had normal vaginal discharge, 5.71% recorded mild vaginal discharge and only 2.86% recorded moderate or no change.

### 2. Burning and/or vaginal pain

After 90 days, symptoms assessment (burning and/or vaginal pain) was performed for 96 patients and 28.13% found that the burning/vaginal pain had disappeared, 70,83% recorded favorable changes in the symptoms (a light or moderate sensation) and only 1.04% had the same symptoms as before treatment. At the second visit, out of 93 participants evaluated, 68.82% reported no vaginal pain, 24.73% reported a mild pain, and 6.45% reported a moderate pain. At the final visit, 96.55% of all evaluated participants reported no vaginal pain/burning, and 1.15% reported mild, moderate, or similar symptoms to pre-treatment. Figure 3 describes vaginal discharge evaluation performed at Visit 1, Visit 2 and Visit 3.

**Figure 3.**
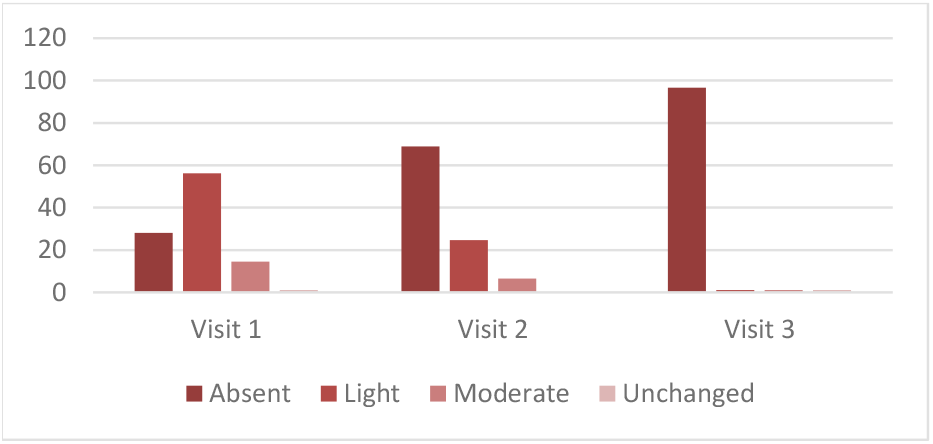
Burning/vaginal pain evaluation at every visit

A comparison between participants treated with monotherapy vs participants treated with Cerviron^®^ as adjuvant treatment was performed, as shown in figure 4. When applied as monotherapy, 96.30% participants reported no vaginal pain or burning, and 1.85% had mild or moderate symptoms after 90 days of treatment, while when using Cerviron^®^ in conjunction with antibiotic treatment, a percent of 94.12% reported no vaginal pain/burning, and only 2.94% had pre-treatment symptoms.

**Figure 4.**
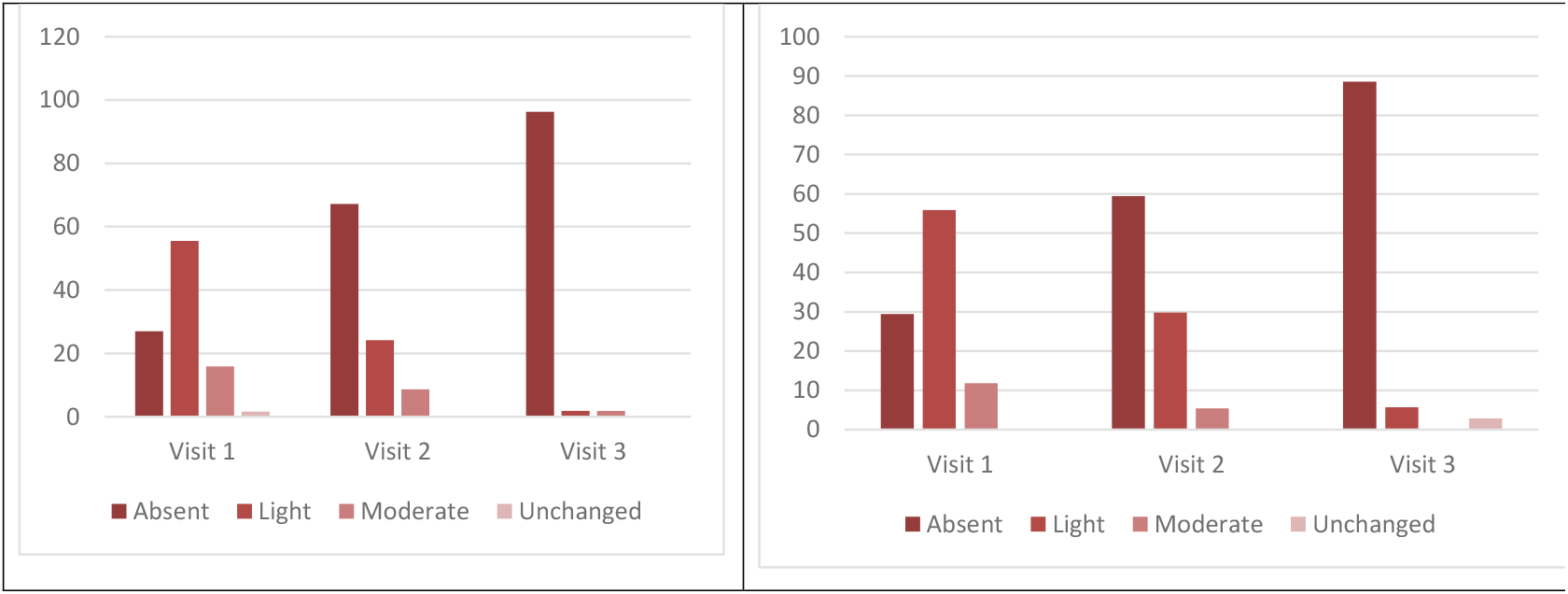
Performance of Cerviron® as monotherapy (left) vs efficacy of Cerviron® ovules as adjuvant treatment (right) in burning/vaginal pain relief

### 3. Vaginal irritations

Ninety-two patients were evaluated for vaginal irritation at the first visit. After 90 days of treatment, 42.39% reported resolution of symptoms, while symptoms were mild in 42.39% and moderate in 15.22%. At Visit 2, 72.63% of the 95 participants reported no vaginal irritation, 22.11% rated the symptoms as mild, and 4.21% rated them as moderate. Significant improvement was seen at the last visit after 3-months treatment with Cerviron^®^ ovules, with 96.55% of the 87 women recording no vaginal irritation. Only 2.30% had mild inflammation and 1.15% had residual pre-treatment symptoms, as seen in figure 5.

**Figure 5.**
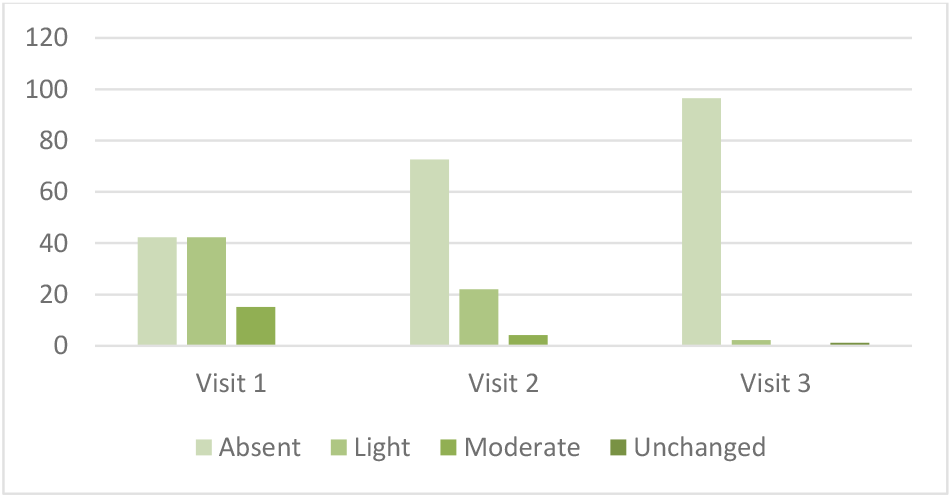
Vaginal irritation evaluation at every visit

#### Monotherapy

After 60 days, 67.80% participants reported no vaginal irritation, 25.42% participants rated the symptoms as mild, and 5.08% participants rated them as moderate. Significant improvement was seen at the last visit after 3 months of Cerviron^®^ monotherapy, as follows: in 54 participants evaluated, 98.15% reported no vaginal irritation and 1.85% reported only a mild irritation.

#### Adjuvant treatment

Thirty-four patients were evaluated for vaginal irritation at the first visit, with 47.06% reporting this symptom absent, while 14.71% and 35.29% reported the symptom as mild or moderate. At visit 2, 78.38% of the 37 participants reported no vaginal irritation, 16.22% rated the symptoms as mild, and 2.70% rated them as moderate. A significant improvement was seen at the final visit after 3 months of treatment when 91.18% of the 34 participants reported no vaginal irritation. In a small percent of 2.94% the irritation remained unchanged. Figure 6 shows the performance of Cerviron^®^ ovules as monotherapy (left) vs Cerviron^®^ ovules as adjuvant treatment (right) in vaginal irritation relief.

**Figure 6.**
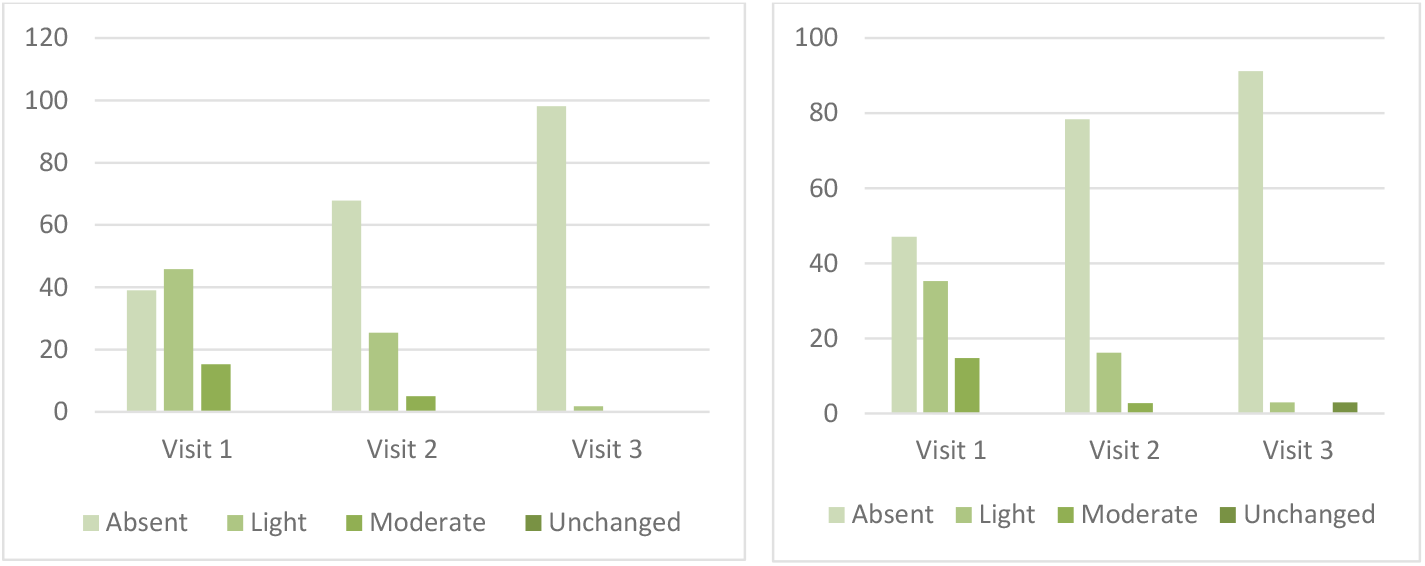
Performance of Cerviron® ovules as monotherapy (left) vs Cerviron® ovules as adjuvant treatment (right) in vaginal irritation relief

### 4. Abnormal smell

This symptom assessment was evaluated for 89 patients before the treatment initiation and the results are presented below: 55.06% had no abnormal smell, 29.21% had a slightly abnormal smell, and 14.61% a moderate abnormal smell. After 30 days, 83 women were evaluated and 77.11% cleared the symptoms, while 18.07% had slight smell and 4.82% had moderate odors. After three months, 81 patients were examined and 96.30% had no abnormal smell and 1.23% rated their symptoms as mild, moderate, or unchanged.

#### Monotherapy

The evaluation of the presence of abnormal smell was rated for 58 patients in the first visit, as follows: 50.00% normal smell, 32.76% had light abnormal smell, and 15.52% moderate abnormal smell. At second visit, 78.00% participants recorded a normal smell. For 18.00% the abnormal smell was lightly present or in moderation in 4.00% participants. After treatment with Cerviron^®^ as monotherapy, all the 50 participants returned to normal. When using Cerviron^®^ in conjunction with antimicrobial treatment, 87.50% from 32 patients evaluated have had a normal smell, while 3.13% presented light, moderate or unchanged abnormal smell, as shown in Figure 8.

**Figure 7.**
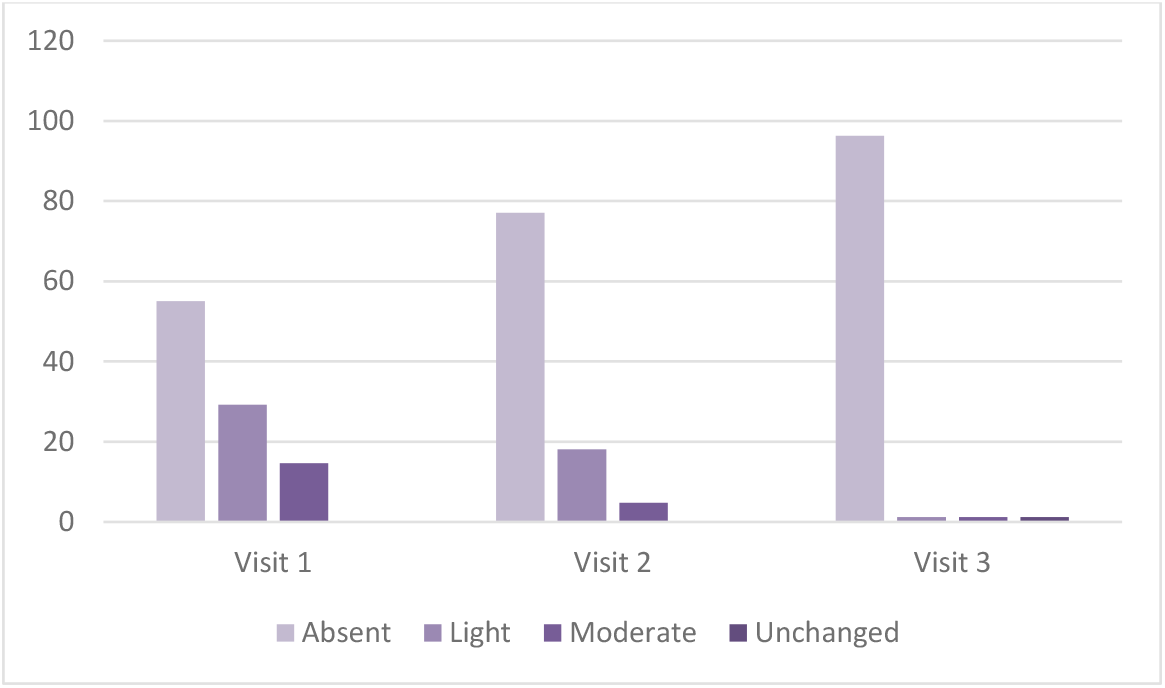
Abnormal smell evaluation at every visit

**Figure 8.**
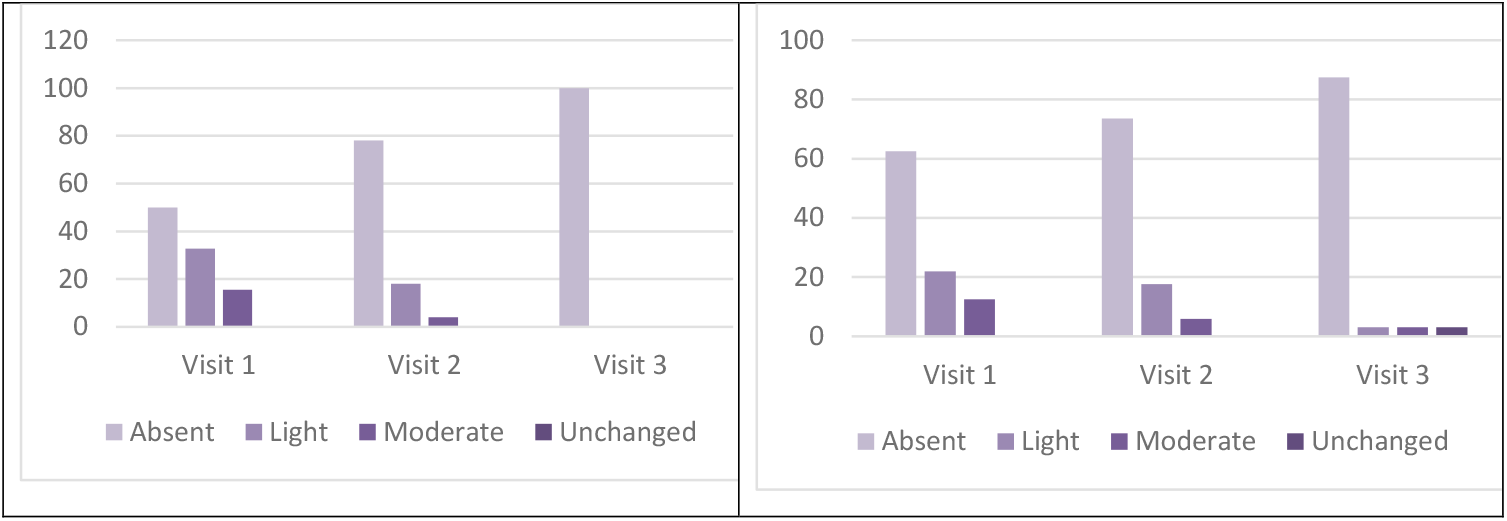
Performance of Cerviron® ovules as monotherapy (left) vs Cerviron® ovules as adjuvant treatment (right) in abnormal smell changes

### 5. Changes in the normal pH level

For changes in the pH level, eighty-three participants were evaluated at visit 1. At visit 3, the following data was recorded: normal pH in 42.17% of the total number of participants, slightly modified in 8.43%, and abnormal in 3.61%.

The vagina has a dynamic microbial ecosystem with varying vaginal pH levels. An imbalance in that ecosystem can alter the vaginal pH and upscale to the point of causing medical attention. It is observed that after following a 3 months’ treatment with Cerviron^®^ 42.17% presented a normal pH, 8.43% of the total participants presented a slightly modified pH, and an abnormal pH was recorded in 3.61%. The normal vaginal pH level (between 3.8 – 4.5) is very important for its protective role in blocking yeast and bacteria multiplication. Thus, the supportive role of the medical device in preventing reinfection is proven on the basis of its effective role in correction of the unbalanced vaginal pH.

### 6. Degree of satisfaction after using Cerviron^®^ ovules

Regarding the patient degree of satisfaction, the largest population of participants (76.43%) indicated that they were very satisfied with the medical device, and none of the patients evaluated declared themselves unsatisfied or very unsatisfied (see figure 10).

**Figure 9.**
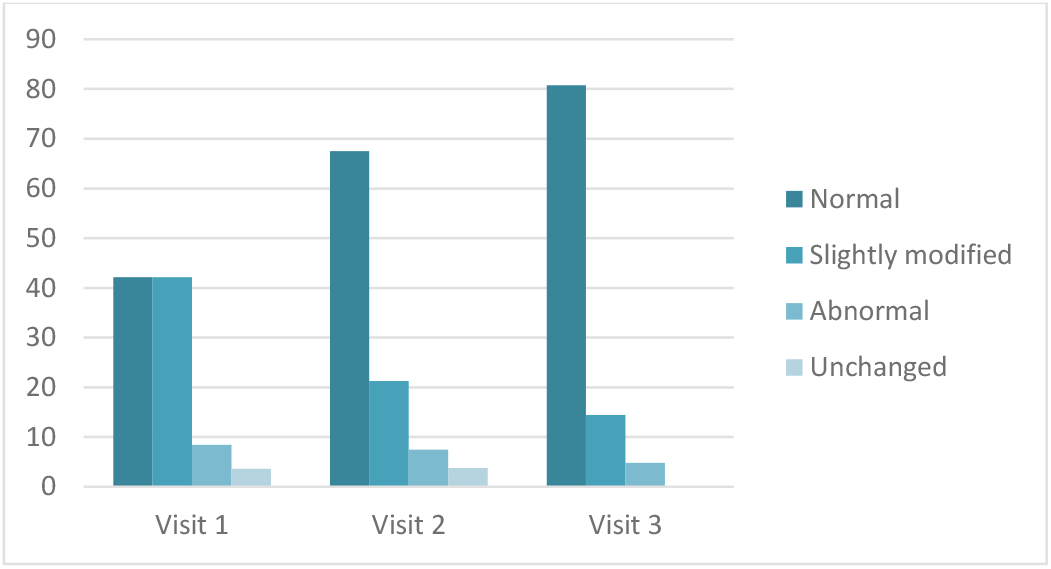
PH Changes between visits

**Figure 10.**
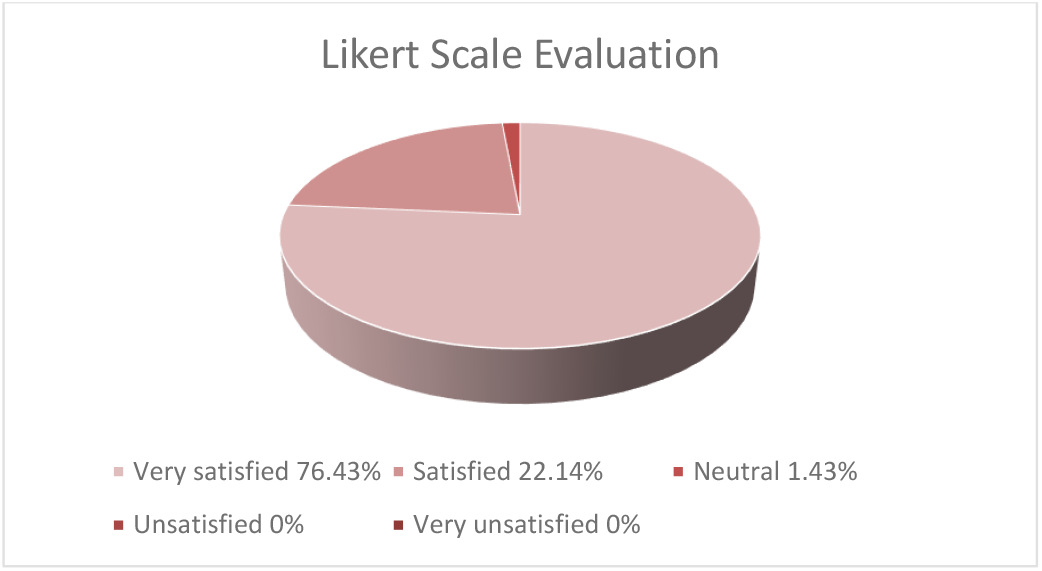
Degree of satisfaction after using Cerviron® ovules

## Discussion

Normally, a healthy vaginal environment maintains a balance between the protective organisms (vaginal Lactobacilli) and other anaerobic and aerobic flora. A typical concentration of the vaginal fluid is maintained by Lactobacilli producing H_2_O_2_ and as such, an acidic vaginal environment. Non-specific vaginitis (also called vaginal dysbiosis) is the disruption of the vaginal microbiome, triggering higher pH values, and specific vaginal symptomatology. Recent studies show that the vaginal dysbiosis affects human papilloma virus acquisition, persistence, and progression to related cervical premalignancy. (18)

Vaginal inflammation is often present in the diagnosis of vaginitis. The present investigation shows that the medical device reduces vaginal inflammation, that is further translated in symptomatology relief (pain and burning reduction). Hence, Cerviron^®^ supportive therapy may be prescribed for 10 or 15 consecutive days. While balancing the vaginal pH, Cerviron^®^ is able to reduce vaginal inflammation and to improve the amount and aspect of vaginal discharge.

A total of 111 patients received Cerviron® vaginal ovules over a treatment period of 3 months and experienced no side effects related to the use of the medical device. Seventy-one participants received Cerviron® ovules as monotherapy and 40 participants used the medical device in combination with other antibiotics. This allowed us to gather more clinical data about possible side effects during different treatment strategies. No side effects have been reported. Hence, Cerviron^®^ can be safely applied in different forms of vaginitis. When added to the antimicrobial therapy, Cerviron^®^ is able to restore the vaginal pH, to reduce inflammation and to reduce the proliferation of bacteria. With a wide spectrum of vaginal symptoms, vaginitis may have important consequences in terms of discomfort and pain, days lost from school or work, sexual functioning, and self-image. As reported by the current guidelines, the most bothersome vaginal symptoms include vulvovaginal itching, burning, irritation, pain, “fishy” vaginal odor, and abnormal vaginal discharge. (19) Distinguishing vaginal from vulvar symptoms is important to direct evaluation and treatment. Vaginitis is correlated with sexually transmitted diseases and other infections of the female genital tract, including HIV as well as with negative pregnancy outcomes. Many women with chronic vulval and vaginal symptoms perform self-medication with topical agents (antibiotics, antifungal agents, corticosteroids, and combinations) and systemic treatments that may mask or exacerbate the symptoms, making diagnosis difficult. In cases that are refractory to treatment, the diagnosis should be reconsidered. (20)

Real-world evidence studies are post-marketing studies bringing valuable information related to the medical devices’ safety and performance profiling and a broader understanding of the practice pattern and the clinical outcomes. The rationale of the study was aimed at capturing safety data in a broader, more heterogenous population. Cerviron^®^ vaginal ovules have been used with success in the treatment of acute and chronic vulvovaginitis of mechanical etiology, and in cervical lesions of mechanical origin, but with a limited number of study participants (NCT04735705 and NCT04735718). (21,22)

## Conclusions

Considering the re the results of the current study, it was concluded that the medical device Cerviron^®^ ovules is safe to use with no adverse effects reported.

The results show that most of the patients reported a decrease or no presence of the above-mentioned symptoms after following a three months’ treatment. No persistence or worsening of symptoms /clinical signs were noted in any participant. The presence of vaginal symptoms has improved statistically significant after 90 days.\. In terms of the degree of satisfaction offered, participants in the largest proportion (around 76.3%) said they were very satisfied with this medical device and 22.14% were satisfied. Only 1.43% of the total number of participants declared themselves neutral in terms of satisfaction. As a general conclusion, it seems that Cerviron^®^ ovules is effective both as a standalone treatment, and also as an adjuvant treatment when added to anti-infectious therapeutic schemas. The analysis in symptomology relief of patients treated for vaginitis show that the symptoms have been absent after the treatment in both cases, with similar success percentages.

Further studies need to be planned to confirm the performance and tolerability profile of Cerviron^®^ in long-term use (prevention of vaginitis relapses) and in patients with mixed aerobic infection and its tolerability in pregnancy.

## Data Availability

All data produced in the present study are available upon reasonable request to the authors.

